# SGLT2 inhibitors and the risk of diabetic ketoacidosis among adults with Type 2 Diabetes: A systematic review and meta-analysis

**DOI:** 10.1101/2021.03.17.21253796

**Authors:** Michael Colacci, John Fralick, Ayodele Odutayo, Michael Fralick

**Author notes:** Correspondence: Michael Colacci, MD, St. Michael’s Hospital, 30 Bond Street, M5B 1W8. Study concept and design: All authors. Acquisition of data: All authors. Analysis/interpretation of data: All authors. Drafting of the manuscript: Colacci M, Fralick M. Critical revision of the manuscript: All authors. Statistical analysis: Colacci M, Fralick M, Odatuyo A. Obtained funding: No funding was received for this study. Conflicts of interest: Dr. Colacci, Dr. J Fralick, Dr. Odutayo, and Dr. M Fralick report no conflicts of interest.

## Abstract

**Importance:** The risk of diabetic ketoacidosis (DKA) with sodium-glucose cotransporter-2 (SGLT2) inhibitors is unclear.

**Objective:** To examine the risk of DKA with SGLT2 inhibitors in both observational studies and large clinical trials.

**Data Sources:** Searches of PubMed, EMBASE and CENTRAL (inception to 15 April 2019) without language restrictions; conference proceedings; and reference lists.

**Study Selection:** Randomized controlled trials and observational studies that quantified the rate of diabetic ketoacidosis with an SGLT2 inhibitor in comparison to another diabetes medication or placebo.

**Data Extraction and Synthesis:** Two independent investigators abstracted study data and assessed the quality of evidence. Data were pooled using random effects models with the Hartung-Knapp-Sidik-Jonkman method.

**Main Outcome and Measures:** Absolute event rates and hazard ratios for diabetic ketoacidosis were extracted from each study.

**Results:** Seven randomized trials encompassing 42,375 participants and five cohort studies encompassing 318,636 participants were selected. Among the 7 randomized controlled trials, the absolute rate of DKA among patients randomized to an SGLT2 inhibitor ranged from 0.6 to 2.2 events per 1000 person years. Four randomized trials were included in the meta-analysis, and compared to placebo or comparator medication, SGLT2 inhibitors had a 2.4-fold higher risk of DKA (Relative Risk⍰[RR] =⍰2.46 [95% CI, 1.16-5.21]; I2⍰=⍰0%; P⍰= 0.54). Among the 5 observational studies, the absolute rate of DKA associated with SGLT2 inhibitor use ranged from 0.6 to 4.9 per 1000 person years and a 1.7-fold higher rate of DKA compared to another diabetes medication (RR⍰=⍰1.74 [95% CI, 1.01-2.93]; I2⍰=⍰45%; P⍰= 0.12).

**Conclusions and Relevance:** In adults with type 2 diabetes, SGLT2 inhibitors increase the risk of DKA in both observational studies and large randomized clinical trials.

**Registration:** CRD42019146855

**Funding Source:** None

**KEY MESSAGES:**

‐ Case reports and observational studies have suggested that SGLT2 inhibitors may be associated with an increased risk of DKA, but this finding has not been reproduced in older meta-analyses of randomized controlled trials.
‐ In this systematic review and meta-analysis of randomized trials and observational studies including over 350,000 patients, SGLT2 inhibitors were found to be associated with twice the risk of diabetic ketoacidosis versus placebo or a comparator medication.
‐ Patients receiving an SGLT2 inhibitors should be counselled on this risk and provided with appropriate sick-day medication management.

## INTRODUCTION

Sodium-glucose co-transporter 2 (SGLT2) inhibitors were approved in 2013 for the treatment of adults with type 2 diabetes mellitus.^1^ Unlike most other diabetes medications, they have consistently been shown to reduce the risk of myocardial infarction, cardiovascular mortality, heart failure, and renal failure.^2–5^ SGLT2 inhibitors are recommended as a second-line agent after metformin by most international diabetes associations.^6,7^ The recently published DAPA-HF (Dapagliflozin in Patients with Heart Failure and Reduced Ejection Fraction) trial demonstrated a lower risk of cardiovascular death or hospitalization for heart failure among patients with and without type 2 diabetes who were randomized to an SGLT2 inhibitor as compared with placebo.^8^ Thus, SGLT2 inhibitors may soon be indicated for a wider patient population, which highlights the importance of identifying and quantifying the risk of rare adverse events.^9^

Diabetic ketoacidosis (DKA) is a potential rare adverse event associated with SGLT2 inhibitors in adults with type 2 diabetes.^10^ An increased risk of diabetic ketoacidosis was not detected until widespread use in routine care.^11,12^ These findings were surprising since DKA is typically a complication of type 1 diabetes mellitus rather than type 2 diabetes,^13^ and is usually characterized by marked hyperglycemia and acidemia due to profound insulin deficiency.^13^ By contrast, initial case reports of DKA among patients who received an SGLT2 inhibitor, typically occurred in the absence of severe hyperglycemia (“euglycemic” diabetic ketoacidosis, blood glucose <200 mg/dL).^10^ As case reports accumulated, the US Food and Drug Administration (FDA) released a warning in May 2015 and updated the drug label in December 2015 to list DKA as a potential side-effect of SGLT2 inhibitors.^14^

Since this FDA warning, several observational studies have identified an increased incidence of DKA with SGLT2 inhibitors compared to other diabetes medications when used in routine care.^9,15^ However, three systematic reviews and meta-analyses of randomized controlled trials (RCTs) found that SGLT2 inhibitors were not associated with an increased risk of DKA.^16–18^ There are several reasons why these meta-analyses may not have identified an increased risk of DKA. First, patients included in RCTs are typically healthier than those in routine care.^19^ Second, RCTs are not powered to detect rare adverse events.^19,20^ Third, these meta-analyses do not include the most recently completed RCTs of SGLT2 inhibitors. Accordingly, we conducted a systematic review and meta-analysis of large clinical trials and observational studies to assess the risk of DKA with SGLT2 inhibitors.

## METHODS

### Protocol and Registration

We conducted a systematic review and meta-analysis, according to the Preferred Reporting Items for Systematic Reviews and Meta-Analyses (PRISMA) guidelines and registered it on Prospero.^21^ (CRD42019146855; eMethods in the Supplement).

### Data Sources and Searches

PubMed (Medline), Embase, Central, and Google Scholar were searched from inception until April 15, 2019. Conference abstracts, including unpublished studies, from the American Diabetes Association, Diabetes Canada and European Association for the Study of Diabetes from January 1, 2013 to April 15, 2019 were also reviewed. Additional articles were identified by screening reference lists of articles and contacting study investigators.

Our search strategy is summarized in the eMethods in the Supplement. Keywords included sodium glucose co-transporter-2 inhibitors, canagliflozin, dapagliflozin and empagliflozin. For each database, we used keywords to identify the appropriate controlled vocabulary terms (eg, MeSH headings). We included large RCTs and observational studies (N≥500 patients receiving SGLT2 inhibitors) that quantified the risk of DKA with SGLT2 inhibitors, relative to a control group. Furthermore, the observational studies were required to have a new-user, active comparator study design, where only patients who were newly initiated on either an SGLT2 or comparator medication were included.

### Study Selection

Two authors (M.C and J.F.) independently reviewed all titles and abstracts to determine eligibility; the full text of the article was evaluated if the content was not clear from the abstract. Disagreements in study inclusion were resolved through consensus, and when no consensus was reached (<1% of studies) disagreements were resolved by a third author (M.F.).

### Data Extraction and Quality Assessment

An initial data collection tool was piloted using 5 articles and revised thereafter based on mutual consensus (M.C. and J.F.). The following information was independently extracted from each article by 2 trained investigators (M.C and J.F.): study period, country, study design, inclusion and exclusion criteria, sample size, primary outcome, and study conclusion. We also extracted patient demographics, including age, sex, comorbidities, baseline hemoglobin A1c, and baseline medications (when available). We collected the following information specific to cohort studies: population sample, matching details, outcome type, and method of outcome diagnosis (e.g., International Classification of Diseases [ICD]). For both RCTs and observational studies we extracted hazard ratio estimates for the incidence of DKA. Hazard ratios from observational studies were multivariable adjusted estimates. Finally, we extracted results of the absolute rate of DKA events. We collected the following antihyperglycemic medication information: time from initiation to episode of DKA and number of total antihyperglycemic medications. When relevant information was not included in a publication, including the number of DKA events, we searched for the primary data on www.clinicaltrials.gov or contacted the study authors.

All studies were independently assessed by 2 investigators (J.F. and M.C.) for risk of bias. The Cochrane risk of bias tool was used for randomized controlled trials.^22^ The Newcastle-Ottawa Scale was used for cohort studies.^23^ Disagreements in quality assessment were resolved by consensus.

### Study Outcome

Events of DKA were identified through either clinical adjudication (RCTs) or diagnostic codes (observational studies). Diagnostic codes for diabetic ketoacidosis (ICD-9 250.1, ICD-10 E11.1) generally have high specificity (>90%) and positive predictive value (85%). ^24,25^

### Data Synthesis and Analysis

All statistical analyses were performed using RevMan Version 5.3 (the Nordic Cochrane Centre, the Cochrane Collaboration) and R (Version 3.5.2) independently by two investigators (M.C. and A.O.). To pool study results, we used random-effects models using the method of Hartung-Knapp-Sidik-Jonkman.^26^ Meta-analysis was performed separately for the observational studies and clinical trials. We quantified statistical heterogeneity using the I^2^ test statistic. We evaluated the potential for publication bias with funnel plots for the outcomes. Due to the small number of studies in our meta-analysis, we did not use the Egger test to test for funnel plot asymmetry.^27^

### Pre-Specified Subgroup Analyses

We planned a priori to complete sensitivity analyses subgroup analyses stratified by age, baseline hemoglobin A1c, ethnicity, and duration of follow up. However, the information for these analyses was unavailable, or not available for a sufficient number of studies.

## RESULTS

We identified 21,715 potential articles, and 12 studies met our inclusion criteria (Figure 1). These consisted of 7 RCTs (23,218 patients on SGLT2 inhibitors and 19,057 on a comparator medication or placebo, Table 1A), with 73 DKA events. Likewise, there were 5 cohort studies (159,318 patients on SGLT2 inhibitors and 159,318 patients on a comparator medication, Table 1B) with 262 DKA events.

**Figure 1:**
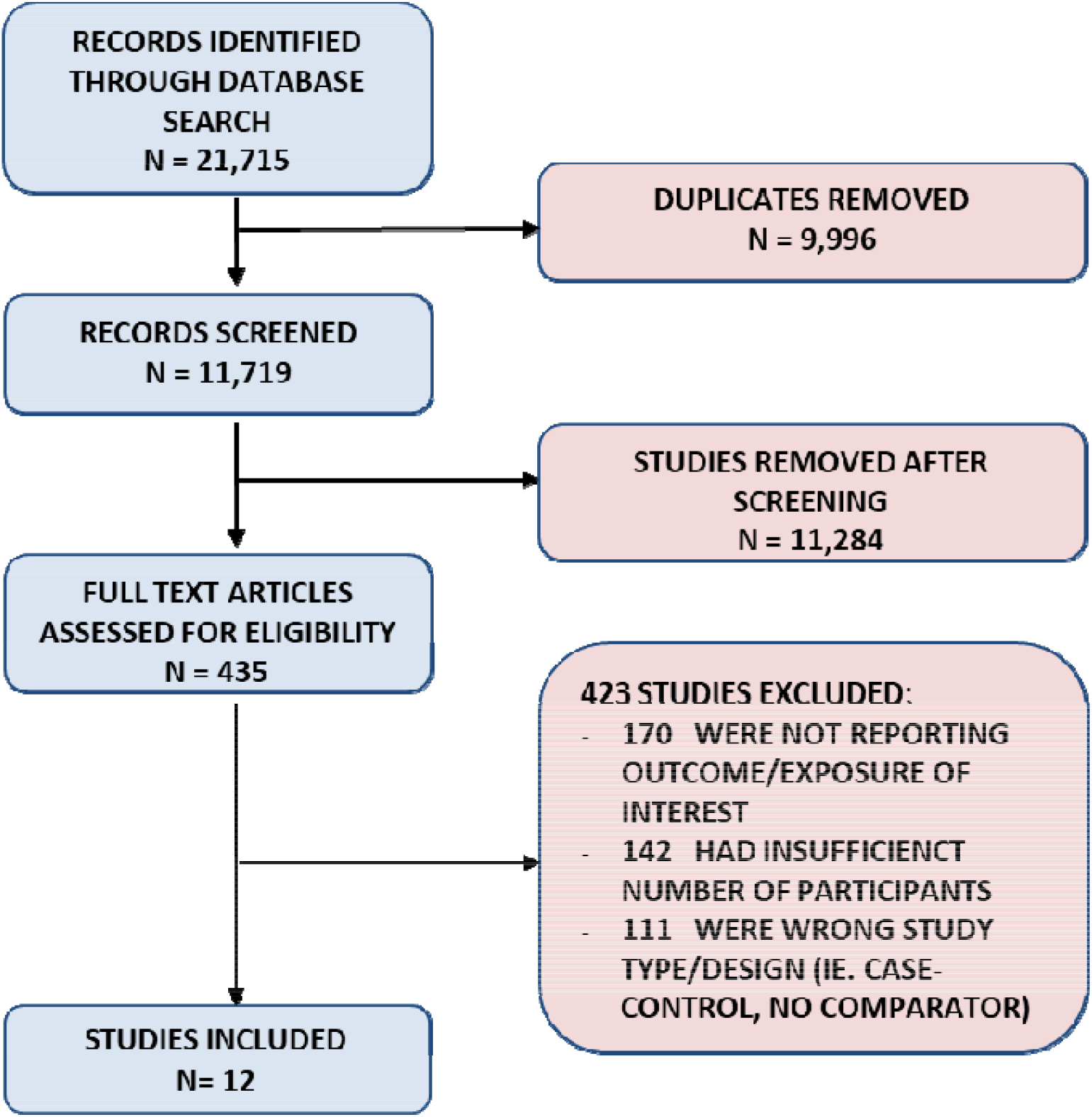
Flow diagram of included studies

**Table 1A:** Study characteristics of included randomized controlled trials

| Author | Country | Study Period | Sample size (N) |  | Study population |  |  | Rate of DKA |  |  |
| --- | --- | --- | --- | --- | --- | --- | --- | --- | --- | --- |
|  |  |  | SGLT2 | Placebo | Age (y) | Female (%) | Insulin use (%) | HR (95% CI) | SGLT2 Per 1000PY | Comparator Per 1000PY |
| Lavalle-Gonzalez F, 2013 | International | 2010-2012 | 735 | 549 | 55.4 | 53.9 | NA | NA | 0* | 1.8* |
| Neal B, 2017 | International | 2009-2017 | 5795 | 4347 | 63.2 | 35.1 | 50.2 | 2.33 (0.76, 7.17) | 0.6 | 0.3 |
| Ridderstrale M, 2018 | International | 2010-2015 | 765 | 780 | 56.2 | 44.0 | 0 | NA | 0 | 0 |
| Wiviott S, 2019 | International | 2013-2018 | 8582 | 8578 | 63.9 | 36.9 | 41.6 | 2.18 (1.10, 4.30) | 0.75* | 0.33* |
| Zinman B, 2015 | International | 2010-2015 | 4687 | 2333 | 63.1 | 28.8 | 48.0 | 1.99 (0.22, 17.80)* | 0.28* | 0.14* |
| Perkovic V, 2019 | International | 2014-2018 | 2202 | 2199 | 62.9 | 34.6 | 65.9 | 10.8 (1.39, 83.65) | 2.2 | 0.2 |
| Tikkanen I, 2015 | International | 2011-2012 | 552 | 271 | 60.3 | 40.8 | NA | NA | 0* | 15.99* |

\*calculated
LEGEND: N: Number, DKA: Diabetic Ketoacidosis, SGLT2: Sodium Glucose Cotransporter-2, y: years, CI: Confidence Interval, PY: Person years, NA: Not available

**Table 1B:** Study characteristics of included observational studies

| Author | Country | Study Period | Sample size (N) |  | Study population |  |  | Rate of DKA |  |  |
| --- | --- | --- | --- | --- | --- | --- | --- | --- | --- | --- |
|  |  |  | SGLT2 | Comparator/Placebo | Age (y) | Female (%) | Insulin use (%) | HR (95% CI) | SGLT2 Per 1000PY | Comparator Per 1000PY |
| Fralick M, 2018 | USA | 2013 - 2015 | 38045 | 38045 (DPP4) | 54.8 | 47.3 | 23.0 | 2.24 (1.40, 3.57) | 4.9 | 2.2 |
| Ueda P, 2018 | Sweden, Denmark | 2013 - 2016 | 17213 | 17213 (GLP1) | 61.0 | 39.0 | 29.0 | 2.14 (1.01, 4.52) | 1.3 | 0.6 |
| Kim Y, 2018 | South Korea | 2013 - 2017 | 56325 | 56325 (DPP4) | 53.1 | 44.9 | 12.6 | 0.96 (0.58, 1.57) | 0.6 | 0.7 |
| Wang Y, 2017 | USA | 2013 - 2014 | 30196 | 30196 (Any) | 54.0 | NA | 11.0 | 1.91 (0.94, 4.11) | NA | NA |
| Patorno E, 2019 | USA | 2014 - 2016 | 17539 | 17539 (DPP4) | 58.7 | 46.3 | NA | 2.16 (1.02, 4.58) | 2.9 | 1.3 |

LEGEND: N: Number, DKA: Diabetic Ketoacidosis, NA: Not available, y: years, CI: Confidence Interval, PY: Person years, USA: United States of America, SGLT2: Sodium-glucose Cotransporter-2, GLP1: Glucagon-Like Peptide-1, DPP4: Dipeptidyl peptidase-4, Any: Any antihyperglycemic medication)

Among the RCTs, the average age of patients randomized to an SGLT2 inhibitor ranged from 55.4 years to 63.9 years, and the percentage of female patients ranged from 28.8% to 53.9% (Table 1A). All RCTs were international studies and industry funded.^2–5,28–30^ The SGLT2 inhibitor used was canagliflozin in three studies, empagliflozin in three studies and dapagliflozin in one study. ^2–5,28–30^ The comparator was placebo in five studies, glimepiride in one study and placebo/sitagliptin in one study.^2–5,28–30^ All RCTs identified diabetic ketoacidosis through clinical adjudication.

In the observational studies, the average age of participants ranged from 53.1 years to 64.0 years, and the percentage of female patients ranged from 39.0% to 47.3% (Table 1B). Three observational studies were conducted in the US, one in South Korea, and one in Scandinavia.^9,15,31–33^ Two of the observational studies were industry funded.^32,33^ The SGLT2 inhibitor used was empagliflozin in one study, any SGLT2 inhibitor in two studies and unspecified in two studies.^9,15,31–33^ For three studies the comparator medication was a DPP4 inhibitor, one study used GLP1 agonists and one used any diabetes medication.^9,15,31–33^ All studies used propensity-score matching and identified diabetic ketoacidosis using validated diagnostic codes (ICD-9 or ICD-10).^24^

### Risk of Diabetic Ketoacidosis Reported in Randomized Trials

Four studies had a DKA event in both the SGLT2 inhibitor and comparator group and were included in the meta-analysis. Of these studies, the duration of follow up ranged from 2.6 to 4.2 years. The absolute rate of DKA among patients randomized to an SGLT2 inhibitor ranged from 0.6 to 2.2 events per 1000 person years, compared to 0.1 to 0.3 per 1000 person years among patients in the control group. This corresponded to a 2.4-fold higher risk of DKA for people receiving an SGLT2 inhibitor compared with placebo/comparator medication (RR⍰=⍰2.46 [95% CI, 1.16-5.21]; I^2^⍰=⍰0%; P⍰= 0.54, Figure 2).

**Figure 2.**
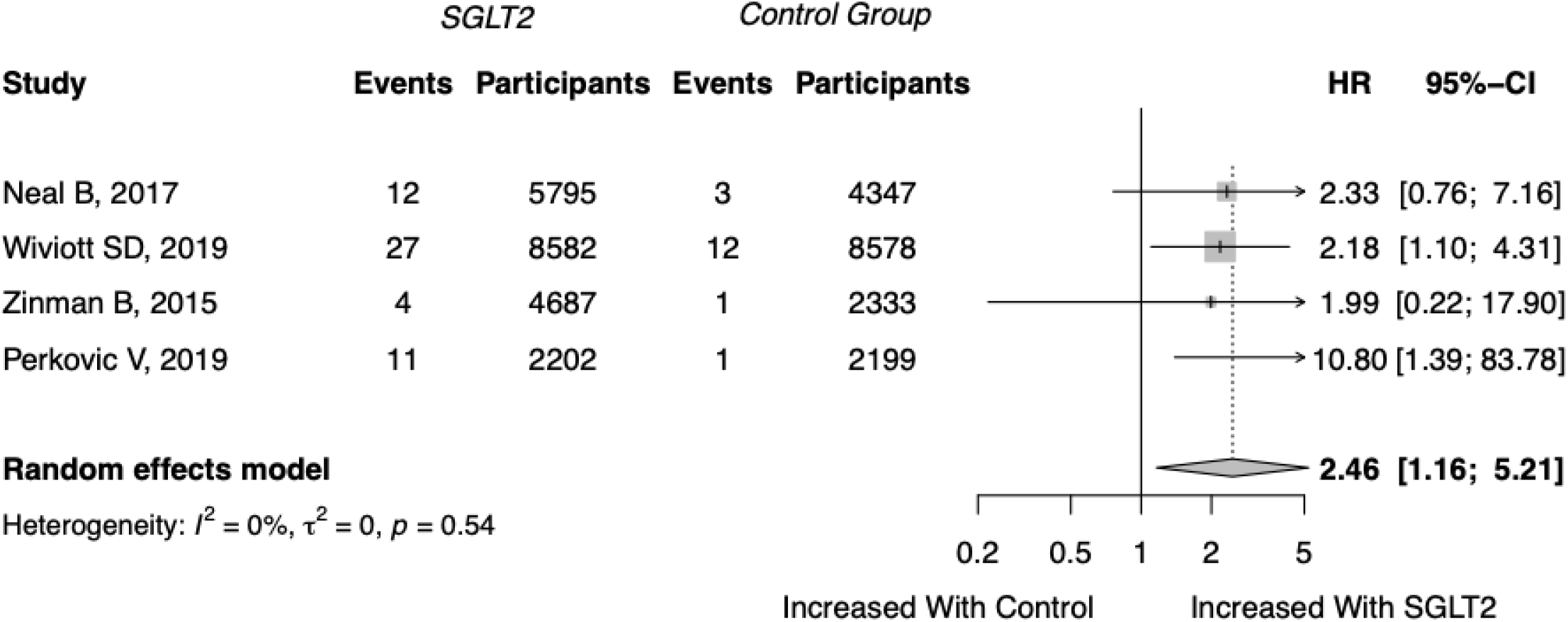
Forest plot of diabetic ketoacidosis risk with sodium-glucose cotransporter-2 (SGLT2) inhibitors vs. comparator in randomized controlled trials Legend: CI: Confidence Interval

### Risk of Diabetic Ketoacidosis Reported in Observational Studies

Median patient follow up among the observational studies ranged from approximately 6 to 12 months. The absolute rate of DKA among patients taking SGLT2 inhibitors in observational studies ranged from 0.6 to 4.9 per 1000 person years compared to 0.6 to 2.2 per 1000 person years among patients who received an active comparator. This corresponded to a 1.7-fold higher risk of DKA for people receiving an SGLT2 inhibitor compared to the active comparator (RR⍰=⍰1.74 [95% CI, 1.07-2.83]; I^2^⍰=⍰45%; P⍰= 0.12, Figure 3).

**Figure 3.**
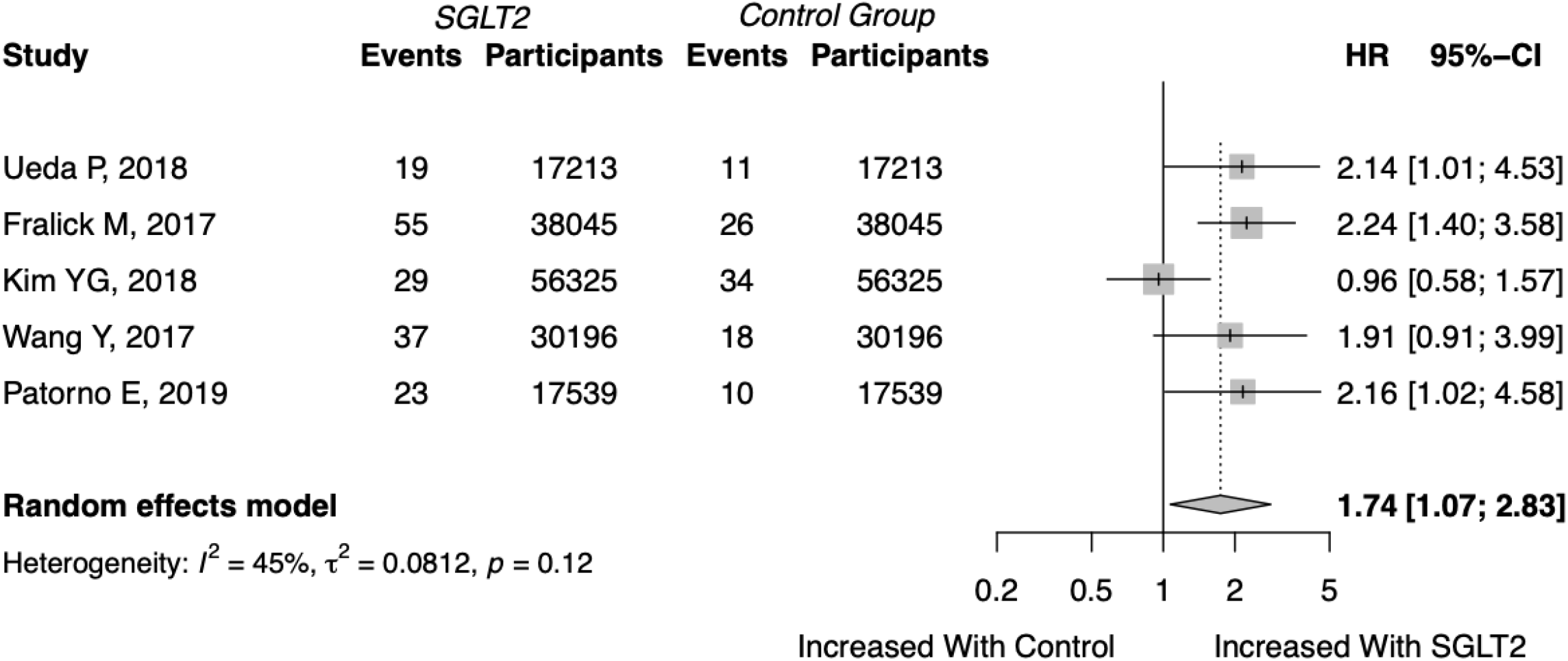
Forest plot of diabetic ketoacidosis risk with sodium-glucose cotransporter-2 (SGLT2) inhibitors vs. comparator in observational studies Legend: CI: Confidence Interval

### Risk of Bias

We evaluated each randomized trial using the Cochrane Risk of Bias Tool (eTable 1 in the Supplement). All RCTs were found to have a low risk of bias. We evaluated each observational study using the Newcastle-Ottawa Scale quality assessment criteria (eTable 2 in the Supplement). Of 9 possible points, the median score for cohort studies was 8 (range, 7-9).

### Assessment of Publication Bias

Visual inspection of the funnel plot showed minimal asymmetry (eFigure 1 in the Supplement).

## DISCUSSION

In this systematic review and meta-analysis that included over 360,000 adults with type 2 diabetes, SGLT2 inhibitors were associated with an increased risk of DKA. This finding was consistent regardless of study design (i.e., randomized trial versus cohort study) or comparator group (i.e., placebo versus active comparator). The risk was approximately 2-fold higher than with placebo/comparator medications, though the absolute rate was low (i.e., 0.6 - 4.9 events per 1000 person years).

Three prior meta-analyses have assessed the risk of DKA with SGLT2 inhibitors, however, none included observational studies.^16–18^ One industry-funded meta-analysis included 8 RCTs (N=10,157 patients taking SGLT2 inhibitors) and found that the risk for DKA was similar in adults randomized to SGLT2 inhibitors compared with placebo (OR 1.14 [95% CI 0.45 to 2.88], P = 0.78).^17^ A second study assessed the risk of DKA in 26 RCTs (N=9,452 patients taking SGLT2 inhibitors), and found a lower risk with SGLT2 inhibitors compared to placebo (RR 0.66; [95%⍰CI 0.30 to 1.45], I^2^=0.0%) and incretin treatment (RR 0.43; [95%⍰CI 0.07 to 2.75], I^2^=0.0%), although with wide confidence intervals that included the possibility of a null effect.^16^ The third study (N=14,726 patients taking SGLT2 inhibitors) assessed the risk of DKA among 34 RCTs, and found a higher odds of DKA with SGLT2 inhibitors, albeit with wide confidence intervals which included the possibility of a null effect (OR 1.36; [95% CI 0.39–4.74], P = 0.63, I^2^ = 0%).^18^

In contrast, our study identified an increased risk of DKA among patients receiving SGLT2 inhibitors in both randomized controlled trials and observational studies. There are two principal reasons why our results differ from the aforementioned meta-analyses. First, we restricted our meta-analysis to large clinical trials, as smaller trials are likely to be under-powered to detect the rare event of DKA. Second, our study included two RCTs that were recently published, and thus not included in the previous meta-analyses; both were large cardiovascular outcome trials that identified an increased risk of DKA among patients randomized to an SGLT2 inhibitor.^3,5^

The mechanism by which SGLT2 inhibitors cause DKA remains uncertain. The SGLT2 receptor is primarily expressed in the proximal tubule of the nephron, which does not explain why it would potentially lead to DKA. However, a recent study identified that SGLT2 was also expressed in the glucagon-secreting alpha cells in the pancreas and directly triggered glucagon secretion.^34^ Furthermore, by causing persistent glycosuria, SGLT2 inhibitors induce a catabolic state, which indirectly causes glucagon and catecholamine secretion.^35,36^ Lastly, SGLT2 inhibitors have been shown to decrease insulin secretion by pancreatic beta-cells.^37^ The resulting imbalance in insulin and counterregulatory hormones can result in DKA by promoting lipolysis and ketogenesis, which can be exacerbated by any additional cause of volume depletion or catecholamine excess (e.g, concurrent illness, reduced food/water intake, etc.).^35^

There are several limitations to our study. First, the studies primarily included middle-aged adults in North America and thus it is unknown how our results might apply to other populations. Second, the included studies controlled for baseline confounders, but not time-varying confounders or events that occurred after starting the medication (e.g., surgery). Recent data suggest that patients are particularly prone to DKA during the post-operative period which has led to recommendations to hold SGLT2 inhibitors post-operatively for a minimum of one day and until the patient is eating a normal diet.^38,39^ Third, due to the limited number of available studies we were unable to perform planned secondary analyses to identify patient-level factors associated with DKA, or differences in risk between individual SGLT2 inhibitor molecules (e.g., canagliflozin, empagliflozin and dapagliflozin). Lastly, although five of the 12 studies in our meta-analysis were observational in nature, all had identical study designs, comparator groups, statistical analysis, and outcome definitions

DKA is an important, albeit rare, potentially life-threatening adverse event associated with the use of SGLT2 inhibitors. Our data suggest the relative risk of DKA is approximately two times higher than with other antihyperglycemic medications, and the absolute rate is approximately 1 per 1000 person years. Patients should be informed of this risk and counselled to temporarily hold their SGLT2 inhibitors under specific circumstances (e.g., sick day, perioperatively). Providers should be alert to the possibility of euglycemic DKA in patients taking SGLT2 inhibitors. Future studies are needed to identify those at greatest risk.

## Supporting information

MOOSE Checklist

PRISMA Checklist

PRISMA Protocol

## Data Availability

All data were abstracted from previously reported trials.

## Acknowledgements

We thank Dr. Emily Hughes, Dr. Stephanie Lee, Ushma Purohit, Afsaneh Raissi and Carter Winberg for revisions to earlier versions of the manuscript.

