## Supplementary material for "SGLT2 inhibitors and the risk of diabetic ketoacidosis among adults with Type 2 Diabetes: A systematic review and meta-analysis": MOOSE Checklist

### MOOSE (Meta-analyses Of Observational Studies in Epidemiology) Checklist

A reporting checklist for Authors, Editors, and Reviewers of Meta-analyses of Observational Studies. You must report the page number in your manuscript where you consider each of the items listed in this checklist. If you have not included this information, either revise your manuscript accordingly before submitting or note N/A.

| Reporting Criteria | Reported (Yes/No) | Reported on Page No. |
|---|---|---|
| **Reporting of Background** | | |
| Problem definition | Yes | 5 |
| Hypothesis statement | Yes | 5 |
| Description of Study Outcome(s) | Yes | 4-5 |
| Type of exposure or intervention used | Yes | 4-6 |
| Type of study design used | Yes | 5 |
| Study population | Yes | 5 |
| **Reporting of Search Strategy** | | |
| Qualifications of searchers (eg, librarians and investigators) | Yes | 6 |
| Search strategy, including time period included in the synthesis and keywords | Yes | 6, appendix |
| Effort to include all available studies, including contact with authors | Yes | 6 |
| Databases and registries searched | Yes | 5-6 |
| Search software used, name and version, including special features used (eg, explosion) | Yes | Appendix |
| Use of hand searching (eg, reference lists of obtained articles) | Yes | 6 |
| List of citations located and those excluded, including justification | Yes | 6 |
| Method for addressing articles published in languages other than English | Yes | 5-6 |
| Method of handling abstracts and unpublished studies | Yes | 5-6 |
| Description of any contact with authors | Yes | 6 |
| **Reporting of Methods** | | |
| Description of relevance or appropriateness of studies assembled for assessing the hypothesis to be tested | Yes | 6, appendix |
| Rationale for the selection and coding of data (eg, sound clinical principles or convenience) | Yes | 6-7 |
| Documentation of how data were classified and coded (eg, multiple raters, blinding, and interrater reliability) | Yes | 6-7 |
| Assessment of confounding (eg, comparability of cases and controls in studies where appropriate | Yes | 6-7 |

| Reporting Criteria | Reported (Yes/No) | Reported on Page No. |
|---|---|---|
| Assessment of study quality, including blinding of quality assessors; stratification or regression on possible predictors of study results | Yes | 7 |
| Assessment of heterogeneity | Yes | 8 |
| Description of statistical methods (eg, complete description of fixed or random effects models, justification of whether the chosen models account for predictors of study results, dose-response models, or cumulative meta-analysis) in sufficient detail to be replicated | Yes | 8 |
| Provision of appropriate tables and graphics | Yes | 8 |
| **Reporting of Results** | | |
| Table giving descriptive information for each study included | Yes | Table 1, appen |
| Results of sensitivity testing (eg, subgroup analysis) | Yes | 13 |
| Indication of statistical uncertainty of findings | Yes | 10 |
| **Reporting of Discussion** | | |
| Quantitative assessment of bias (eg, publication bias) | Yes | 11, appendix |
| Justification for exclusion (eg, exclusion of non–English-language citations) | Yes | Figure 1 |
| Assessment of quality of included studies | Yes | 11 |
| **Reporting of Conclusions** | | |
| Consideration of alternative explanations for observed results | Yes | 13 |
| Generalization of the conclusions (ie, appropriate for the data presented and within the domain of the literature review) | Yes | 14 |
| Guidelines for future research | Yes | 14 |
| Disclosure of funding source | Yes | 9 |

**Once you have completed this checklist, please save a copy and upload it as part of your submission. DO NOT include this checklist as part of the main manuscript document. It must be uploaded as a separate file.**
