## Supplementary material for "SGLT2 inhibitors and the risk of diabetic ketoacidosis among adults with Type 2 Diabetes: A systematic review and meta-analysis": PRISMA Protocol

PRISMA-P Research Protocol – June 8, 2020

| Title |  | SGLT2 inhibitors and the risk of diabetic ketoacidosis: A systematic review and meta-analysis |
| --- | --- | --- |
| Identification |  | Protocol of a systematic review and meta-analysis |
| Registration |  | PROSPERO: CRD42019146855 |
| Authors | Contact: | Michael Colacci   University of Toronto, Department of Medicine  190 Elizabeth St, Toronto, ON, Canada  Mike Fralick   Staff Internist, Sinai Health System  Assistant Professor, University of Toronto  Research Fellow, Division of Pharmacoepidemiology and Pharmacoeconomics, Brigham and Women’s Hospital, Harvard University |
|  | Contributions | Search development: MC, MF Abstract review: MC, JF  Manuscript Review and Data Collection: MC, JF, MF  Manuscript Preparation: MC, MF, JF, AO  Risk of Bias Assessment: MC, MF, JF  Statistical Analysis: MC, AO, MF  Manuscript Revision: MC, JF, AO, MF |
| Support | Sources: | None |
|  | Sponsor: | None |
|  | Role of sponsor: | None |
| Introduction | Rationale: | Sodium Glucose Co-transporter-2 (SGLT2) Inhibitors have been associated with an increased risk of diabetic ketoacidosis (DKA). However, the relative risk of DKA in comparison to other commonly used diabetes medications is not well known. |
|  | Objectives: | We will review the available literature comparing the risk of diabetic ketoacidosis between an SGLT2 inhibitor and a comparator diabetes medication, and quantify the relative risk of diabetic ketoacidosis. |
| Methods | Eligibility Criteria: | All studies must be   1. Studies in humans 2. Time period: Database inception - present 3. Comparative studies (randomized trials or observational studies) 4. Provide the absolute risk of diabetic ketoacidosis among patients taking an SGLT2 inhibitor AND the absolute risk of diabetic ketoacidosis among patient taking a different diabetes medication, in the same population |
|  | Information sources: | 1. OVID (MEDLINE) 2. OVID (EMBASE) 3. Google Scholar 4. Screen reference lists of articles identified   Grey Literature:   1. American Diabetes Association conference abstracts from (Jan 2013-present) 2. Diabetes Canada Conference (Jan 2013-Present) 3. European Association for the Study of Diabetes (Jan 2013 – present) |
|  | Search Strategy: | 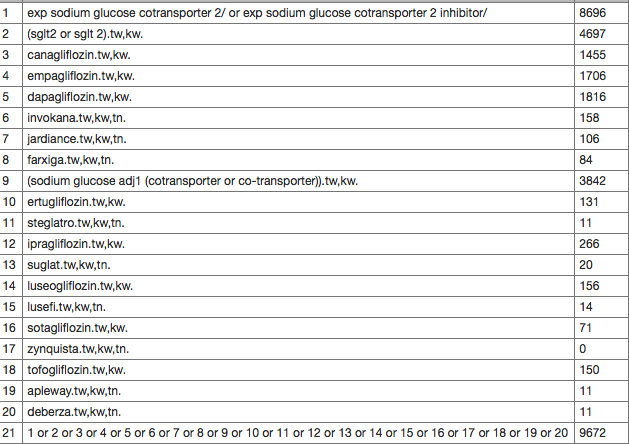 |
|  | Study records: Data management | Covidence |
|  | Selection process | Screening, review and inclusion in meta-analysis will be completed independently by two authors (MC and JF).  Disagreements in study inclusion will be resolved through consensus, and, when no consensus is reached, it will be resolved by a third author (MF). |
|  | Data collection process | Data will be extracted independently by MC and JF |
|  | Data items | Study authorship, year of publication, study period, country, language, study design, inclusion and exclusion criteria, number of patients in SGLT2 and comparator group, absolute risk of DKA in patients taking SGLT2 inhibitors, absolute risk of DKA in patients taking comparator diabetes medication, method of validation (ie. chart review, ICD-9/10 code, etc), measure of quality of study, primary objective of study, study conclusion, patient demographics (age, sex) |
|  | Outcomes and prioritization | 1. Absolute risk of DKA in patients taking SGLT2 inhibitors 2. Absolute risk of DKA in patients taking comparator diabetes medication 3. Unadjusted risk estimates, adjusted risk estimates and the respective 95% confidence intervals of risk of DKA between SGLT2 and comparator medication 4. Baseline hemoglobin A1c in patients taking SGLT2 inhibitors and patients taking the comparator diabetes medication |
|  | Risk of bias in individual studies | Quality Assessment: Newcastle Ottawa Scale for cohort and case-control studies. Cochrane Risk of Bias for randomized trials |
|  | Data synthesis | All statistical analyses will be performed using RevMan Version 5.3 (the Nordic Cochrane Centre, the Cochrane Collaboration) and R (Version 3.5.2) independently by two investigators (M.C. and A.O.). To pool study results, we will use random-effects models using the method of Hartung-Knapp-Sidik-Jonkman. Meta-analysis will be performed separately for the observational studies and clinical trials. We will quantify statistical heterogeneity using the I2 test statistic. We will evaluate the potential for publication bias with funnel plots for the outcomes.  To assess the robustness of our results, we will perform additional analyses stratified by the Newcastle-Ottawa Scale score (lower quality versus higher quality) and restricted to only those studies that provided adjusted risk estimates.  Additional sub-group analysis will be completed on studies in patients from Asia versus those done on patients not from Asia. |
|  | Meta-bias(es) | Publication bias will be evaluated using funnel plots for the outcomes and the Egger test. |
